# Step-downs reduce workers’ compensation payments to encourage return to work. Are they effective?

**DOI:** 10.1101/19012286

**Authors:** Tyler J Lane, Luke R Sheehan, Shannon E Gray, Dianne Beck, Alex Collie

## Abstract

**Objective:** To determine whether step-downs, which cut the rate of compensation paid to injured workers after they have been on benefits for several months, are effective as a return to work incentive.

**Methods:** We aggregated administrative claims data from seven Australian workers’ compensation systems to calculate weekly scheme exit rates, a proxy for return to work. Jurisdictions were further subdivided into four injury subgroups: fractures, musculoskeletal, mental health, and other trauma. The effect of step-downs on scheme exit was tested using a regression discontinuity design. Results were pooled into meta-analyses to calculate combined effects and the proportion of variance attributable to heterogeneity.

**Results:** The combined effect of step-downs was a 0.86 percentage point (95% CI -1.45 to -0.27) reduction in the exit rate, with significant heterogeneity between jurisdictions (*I*^2^ = 68%, *p* = .003). Neither timing nor magnitude of step-downs was a significant moderator of effects. Within injury subgroups, only fractures had a significant combined effect (-0.84, 95% CI -1.61 to -0.07). Sensitivity analysis indicated potential effects within mental health and musculoskeletal conditions as well.

**Conclusions:** The results suggest some workers’ compensation recipients anticipate step-downs and exit the system early to avoid the reduction in income. However, the effects were small and suggest step-downs have marginal practical significance. We conclude that step-downs are generally ineffective as a return to work policy initiative.

**Key messages:** *What is already known about this subject?:* A number of workers’ compensation systems around the world reduce payments to injured workers after they have been in the system for several months. In Australia, where each state, territory, and Commonwealth system employs step-downs, the stated policy objective is to increase the rate of return to work through financial incentives. However, there is little empirical evidence to either support or reject this claim.

*What are the new findings?:* The rate at which claimants exited workers’ compensation systems increased ahead of step-downs taking effect, suggesting an anticipatory effect. However, the effect was relatively small, changing the exit rate by less than a percentage point overall, with substantial heterogeneity between systems.

*How might this impact on policy or clinical practice in the foreseeable future?:* While statistically significant, the findings suggest that step-downs provide workers’ compensation claimants little incentive to return to work. Policymakers may need to reconsider step-downs as a component of scheme design, or justify them according to their original purpose, which was to save costs.

## Introduction

Step-downs reduce the rate of income replacement paid to injured workers after they have been on benefits for a period of several months. They are found in a number of workers’ compensation systems around the world, including several in Europe (Andorra, Croatia, Slovakia, Sweden), Africa (Ethiopia, Republic of Congo, São Tomé and Príncipe, Zimbabwe), Asia (Indonesia, Laos, Singapore, Taiwan), Central America (Belize, Panama), the Middle East (Kuwait, Oman, Qatar), South America (Ecuador),^1^ and one American state (Ohio).^2^ Unique among these is Australia, where each of its nine major workers’ compensation systems implements step-downs.^3^

Step-downs are promoted as an incentive for claimants to return to work.^4–6^ However, there is little direct empirical evidence to support this claim,^7,8^ and that which exists is generally inconclusive.^6,9^ It also contrasts with the original purpose of step-downs when introduced across Australia in the 1980s and 90s, which was to reign in the rising cost of employers’ insurance premiums.^7^ Nevertheless, evidence that more generous benefits increase time off work indicates that an incentivising effect is plausible.^4^

We test whether step-downs increase the rate at which claimants exit workers’ compensation, and moderating effects of their timing and magnitude. Building on evidence that effects of benefit generosity vary by injury,^10^ we also tested effects in claims for fractures, mental health conditions, musculoskeletal conditions, and other trauma subgroups.

## Methods

Study questions and analyses were pre-registered with the Open Science Framework.^11^ We reproduce the analytical approach here and note any deviations.

### Step-downs in Australia

Australia’s six states, two territories, and Commonwealth government have their own workers’ compensation system for injured workers, which cover 94% of the workforce.^12^ Each scheme is cause-based, meaning benefits are contingent on attribution of the condition, whether an acute injury or gradual onset disease (collectively referred to as “injury” in this paper), to employment.^13^ There are considerable differences in overarching policy settings, including whether the scheme allows common law claims, is publicly or privately underwritten, and generosity of benefits.^3^ While each system employs step-downs, they vary in both timing and magnitude, as illustrated in Table 1.

**Table 1.** Wage replacement rate by jurisdiction and time on workers’ compensation benefits, up to 104 weeks; rate changes and values indicated with heatmap. Data derived from the Comparison of Workers’ Compensation Arrangements in Australia and New Zealand reports.

| Jurisdiction | Date range | Nominal caps* | Compensation rate based on weeks in the system† |  |  |  |  |
| --- | --- | --- | --- | --- | --- | --- | --- |
|  |  |  | 0-12 | 13-25 | 26-45 | 46-77 | 78-104 |
| New South Wales | Oct 2012 to Jun 2015 | Maximum | 95% | 80% | 80% | 80% | 80% |
| Victoria | Apr 2010 to Jun 2015 | Maximum | 95% | 80% | 80% | 80% | 80% |
| Queensland (NWE) | Jul 2009 to Jun 2015 | Maximum | 85% | 85% | 75% | 75% | 75% |
| Queensland (QOTE) | Jul 2009 to Jun 2015 | Maximum | 80% | 80% | 70% | 70% | 70% |
| Western Australia (no industrial agreement) | Jul 2009 to Jun 2015 | Maximum | 100% | 85% | 85% | 85% | 85% |
| Western Australia (industrial agreement)§ | Jul 2009 to Jun 2015 | Maximum | 100% | 100% | 100% | 100% | 100% |
| South Australia | Jul 2009 to Jun 2014 | Maximum | 100% | 90% | 80% | 80% | 80% |
| Tasmania | Jul 2010 to Jun 2015 | Minimum | 100% | 100% | 90% | 90% | 80% |
| Northern Territory | Jul 2009 to Jun 2015 | Maximum (step-down only) | 100% | 100% | 90% | 90% | 90% |
| Australian Capital Territory | Jul 2009 to Jun 2015 | Maximum & minimum | 100% | 100% | 65% | 65% | 65% |
| Commonwealth Comcare | Jul 2009 to Jun 2015 | Maximum & minimum | 100% | 100% | 100% | 75% | 75% |

\* Nominal caps are indexed to state average earnings and change annually or in some cases more frequently. We have chosen to denote only whether such caps exist in the relevant time frames. † Step-down timing is the same for all claims, though step-down rates can vary based on a number of characteristics including pre-injury earnings amount, industrial awards, enterprise agreements, and number of dependents. § Regular salary plus overtime, bonuses, and any allowances paid only for first 13 weeks, after which they are no longer paid under Western Australia's industrial agreement arrangements.

Most of these systems have wage replacement caps that set a nominal maximum on what claimants may earn, and a few have minimums. In Queensland, claimants with an industrial agreement, which is a certified specification of industrial matters between employees and employers, are initially compensated at the greater of 85% their Normal Weekly Earnings (NWE, based on individual pre-injury earnings) or the industrial instrument, which at 26 weeks steps-down to the greater of 75% NWE or 70% Queensland Ordinary Time Earnings (QOTE, based on state mean earnings). Claimants not under an industrial instrument are initially compensated at the greater of 85% NWE or 80% QOTE, and step-down to the greater of 75% NWE or 70% QOTE. In the Northern Territory, step-downs are the greater of 1) 75% of weekly earnings up to a maximum nominal cap, or 2) the lesser of a flat rate plus additional income for each dependent or 90% of NWE. In Western Australia, claimants with an industrial agreement are not subject to step-downs and are compensated at 100% of their regular earnings throughout the life of the claim. However, overtime, bonuses, and allowances are compensated up to 13 weeks but not afterwards,^14^ meaning workers who rely on these extra sources of income are effectively subject to step-downs, though of varying magnitudes. Step-down rates are higher in Tasmania and Comcare (the Commonwealth system) if the claimant is back at work in some form of partial capacity.^4,14^ In these cases, the magnitude of initial and step-down compensation rates vary, though timings remained the same.

In Victoria and to a lesser extent New South Wales, claimants from unionised industries often have industrial awards and enterprise agreements that top up payments and can make up any gaps between pre-injury earnings compensation.^7,8^ We were unable to account for these arrangements nor determine what proportion of the population was affected by them.

#### Data

Data were derived from the *National Data Set for Compensation-based Statistics*, an amalgamation of case-level administrative claims data from each system that is compiled by Safe Work Australia.^15^ The pre-registered inclusion criterion restricted eligibility to claims lodged since either July 2009 or the most recent change to step-down arrangements, whichever latest, up to June 2015. For instance, in July 2011 Tasmania altered step-down arrangements via legislative amendment. Only claims lodged afterwards were included in analyses. *Post hoc*, we added several other exclusions:

Claims affected by minimum and maximum caps for weekly payments

Claims lodged after June 2014 in South Australia to allow a one-year buffer with the change in step-downs arrangements implemented in July 2015

Claims exempted from New South Wales’ 2012 legislative amendments, including several occupations (police, paramedics, firefighters, and coalmine workers) and dust diseases^3^ Our outcome – weekly scheme exit rate – was determined using cumulative compensated time off work. While scheme exit does not necessarily entail return to work, and cumulative compensated time off work underestimates the total actual duration, it is nevertheless considered the most accurate measure of time off work when using administrative data.^16^ Several jurisdictions including Victoria and South Australia determine the application of step-downs by counting any calendar week in which there was compensated time loss as a full week,^6,17,18^ whereas Comcare uses cumulative compensated time off work.^4^ In the Victorian and South Australian systems, this means that for some claims, step-downs applied earlier than specified in our analyses.

### Analysis

We calculated scheme exit rates by dividing the number of claims exiting the system each week by those in it at the start of that week. Injury subgroups included fractures, mental health conditions, musculoskeletal conditions, and other trauma. Our pre-registered categorisation separated back and neck from other musculoskeletal conditions, though we have since decided to keep them together as a better conceptual fit. Neurological conditions and all other conditions were excluded due to low numbers.

Data were left-censored at four weeks to exclude residual effects of employer excess, which are the post-injury periods for which employers are responsible for compensation payments. Anecdotal reports suggest claims are less likely to persist only a day or two beyond the employer excess period, tending either to resolve before the employer excess period ends, or to persist for a few days beyond that. In Australia, the longest employer excess periods are 10 working days/two weeks in Victoria/South Australia.^3^ We determined *a priori* that four weeks, while arbitrary, would be sufficient to remove any confounding due to this effect. Exit rates were calculated up to two years, or 104 weeks.

Effects were evaluated with a regression discontinuity design, a powerful quasi-experimental approach that compares outcomes on either side of an arbitrary cut-off. When individuals are unable to control which side of the cut-off they are on, regression discontinuity simulates a randomised control trial.^19,20^ In this study, the assumption was inverted in that we evaluated whether claimants crossed this threshold. This means we cannot treat individuals on either side of the step-down cut-off as exposed or control groups and must interpret the results more cautiously.^21^

We incorporated parametric polynomial estimators to account for non-linear patterns in exit rates, testing up to 10 polynomial terms with separate or same slopes, erring on the side of overfitting,^20^ and selected best-fit models based on the Akaike Information Criterion.^19^ Initially, we tested only separate slopes, but in several cases the fitted lines noticeably diverged from data points near the step-down cut-off. Testing same-slope models as well addressed these issues.

Results are reported as the percentage point change to the exit rate. Coefficients and standard errors were combined into random effects meta-analyses to determine combined effects and the proportion of variance attributable to heterogeneity. We tested the moderating effect of step-down timing and magnitudes using meta-regressions.

Exit rates within a few subgroups became unstable as the number of claims in the system diminished over time. To account for this, we excluded data points where the number of remaining claimants for the week was <500, and did not conduct analyses where there were <20 aggregated data points after the step-down. To illustrate the issue, data points in regression discontinuity plots are coloured black where included and grey where excluded. These exclusions were an ad hoc approach to an analytical problem that only became apparent as we examined the full dataset. As a result, neither Tasmania nor the Australian Capital Territory had sufficient data and were thus excluded from analyses.

### Statistical software and analysis packages

Analyses were conducted in R with RStudio using the following packages: *ggpubr*,^22^ *lubridate*,^23^ *metafor*,^24^ *metaviz*,^25^ *rdd*,^26^ *rddtools*,^27^ *scales*,^28^ *see*,^29^ *tidyverse*,^30^ and *zoo*.^31^ Aggregated data and R code are available on a FigShare repository.^32^

## Results

Data counts with crosstabulations for jurisdiction and injury type are summarised in Table 2. In total there were *N* = 292,060 claim records in this study, the majority of which were musculoskeletal (*N* = 176,297, 60%). The findings were first presented at the Actuaries Institute Injury and Disability Schemes Seminar in Canberra on 11 November 2019.

**Table 2.** Count and row percent of included claims by jurisdiction and injury type

| Jurisdiction | All | Fractures | Mental health | Musculoskeletal | Other trauma | Conditions excluded due to low numbers |  |
| --- | --- | --- | --- | --- | --- | --- | --- |
|  |  |  |  |  |  | Neurological | Other conditions |
| New South Wales | 49 391 | 5693 (11.5%) | 4296 (8.7%) | 29 380 (59.5%) | 7456 (15.1%) | 717 (1.5%) | 1849 (3.7%) |
| Victoria | 75 702 | 8604 (11.4%) | 8256 (10.9%) | 43 729 (57.8%) | 9869 (13.0%) | 1868 (2.5%) | 3376 (4.5%) |
| Queensland | 82 973 | 11 303 (13.6%) | 3584 (4.3%) | 50 722 (61.1%) | 11 533 (13.9%) | 2289 (2.8%) | 3542 (4.3%) |
| Western Australia | 41 967 | 5941 (14.2%) | 1789 (4.3%) | 26 798 (63.9%) | 6008 (14.3%) | 538 (1.3%) | 893 (2.1%) |
| South Australia | 27 055 | 2686 (9.9%) | 3968 (14.7%) | 16 827 (62.2%) | 1694 (6.3%) | 846 (3.1%) | 1034 (3.8%) |
| Northern Territory | 5803 | 939 (16.2%) | 647 (11.1%) | 3158 (54.4%) | 729 (12.6%) | 99 (1.7%) | 231 (4.0%) |
| Comcare | 9169 | 695 (7.6%) | 1778 (19.4%) | 5683 (62.0%) | 390 (4.3%) | 287 (3.1%) | 336 (3.7%) |
| Total | 292 060 | 35 861 (12.2%) | 24 318 (8.3%) | 176 297 (60.4%) | 37 679 (12.9%) | 6644 (2.3%) | 11 261 (3.9%) |

### Step-down impact on scheme exit rates

Across jurisdictions, the combined effect of step-downs on exit rates was a reduction of 0.86-percentage points (95% CI: -1.45 to -0.27). A significant, moderate proportion of the variance in effects was attributable to heterogeneity between jurisdictions (*I*^2^ = 68%, *p* = .003).

Within individual schemes, all significant effects were negative. Three of four significant effects were observed in jurisdictions with the earliest step-downs, occurring at 13 weeks: New South Wales (- 1.65, -3.25 to -0.06), Western Australia (-1.65, -3.07 to -0.23), and South Australia (-2.24, -3.38 to - 1.10). Victoria also had a 13-week step-down, though the effect was non-significant (0.03, -0.88 to 0.95). The only significant effect outside of 13 weeks was in Comcare, where step-downs occur at 45 weeks (-1.29, -2.25 to -0.34). However, meta-regressions found that neither the timing (0.01, -0.08 to 0.09) nor magnitudes (0.02, -0.13 to 0.17) of step-downs significantly moderated the effect on exit rates.

Results are summarised in Figure 1, and regression discontinuities are plotted in Figure 2.

**Figure 1.**
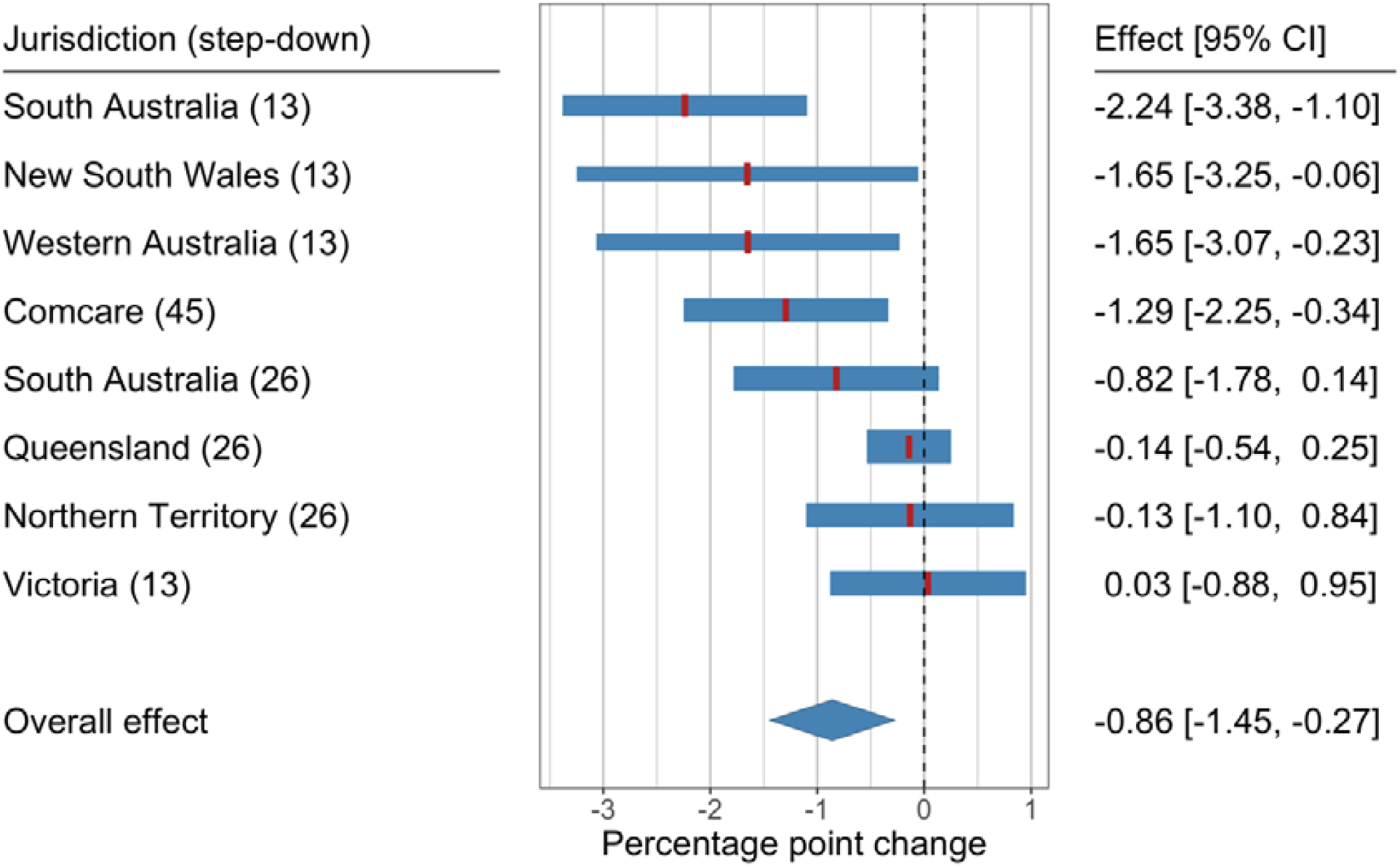
Meta-analysis forest plot of jurisdictional and combined effects of step-downs. Band thickness reflects meta-analytic weight of each jurisdiction

**Figure 2.**
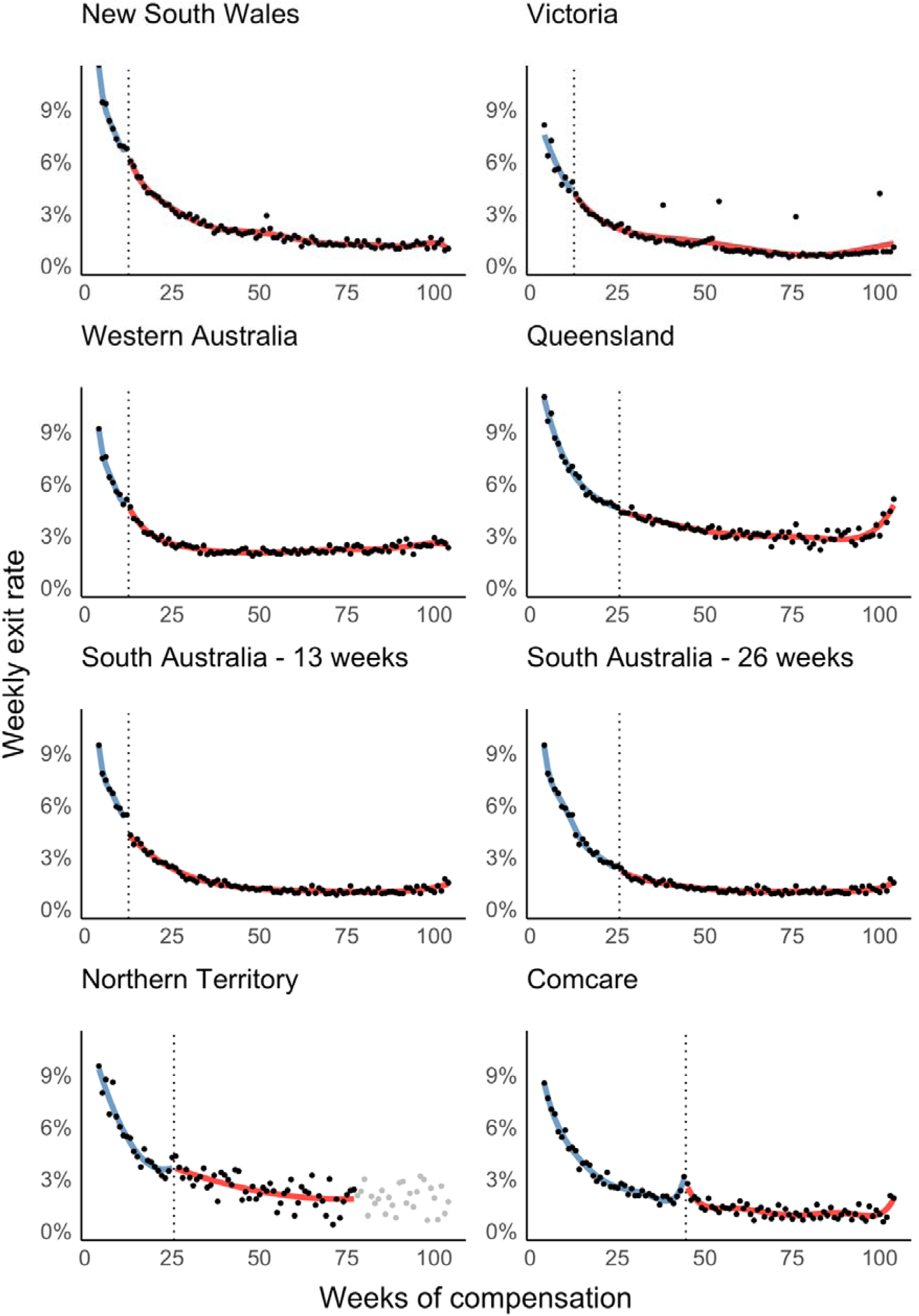
Regression discontinuity plots illustrating impact of step-downs on exit rates by jurisdiction. Grey data points indicate excluded data (<500 denominator cases)

### Sensitivity analysis – confounding from competing incentives

We identified potential confounding from competing scheme incentives such as 10% insurance premiums discounts in New South Wales for employers who return claimants to work within 13 weeks^33^ and bonuses for claims agents in Victoria who keep the rate of claims reaching 13 weeks low.^34^ Other such incentives may exist, though consultation with scheme representatives indicated this information is often confidential as a private arrangement between insurers and employers.

We conducted sensitivity analyses on claimants unaffected by step-downs, which were identified based on pre-injury wages and maximum and minimum wage replacement caps. Significant changes among these claims would be evidence of confounding. Only three jurisdictions (Victoria, Queensland, and Western Australia) had sufficient data for this analysis. Effects were non-significant individually and combined (0.16, -0.50 to 0.82). These results are summarised in Supplementary Figure 1.

### Step-down impact by injury type

Combined effects were significant only among fracture claims (-0.84, -1.61 to -0.07). Heterogeneity between sites was non-significant (*I*^2^ = 25%, *p* = .087). Meta-analyses by injury are summarised in Figure 3, and regression discontinuity plots are presented in Supplementary Figures 2 to 5.

**Figure 3.**
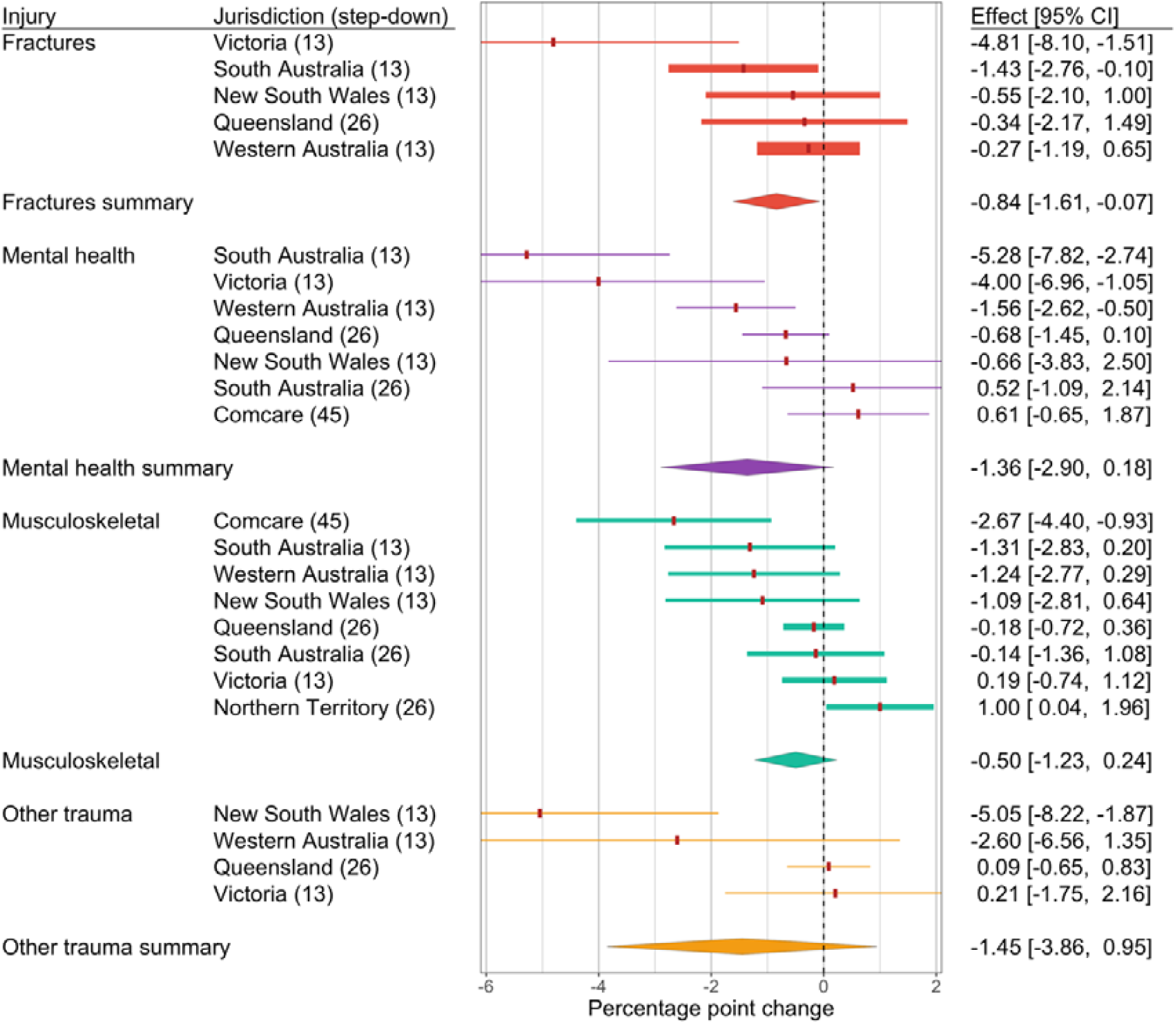
Meta-analysis forest plot of jurisdictional and combined effects of step-downs by injury subgroup. Band thickness reflects meta-analytical weight of each jurisdiction. See Supplementary Figures 2-5 for individual regression discontinuity plots.

### Sensitivity analysis – step-down impact by injury type

While combined effects were non-significant in mental health, musculoskeletal, and other trauma claims, magnitudes were similar across all injury types (-0.50 to -1.45) with considerable overlap in confidence intervals. There were also indications that a single jurisdiction was responsible for attenuation to non-significance in some injuries, such as the lone positive effect among musculoskeletal conditions in the Northern Territory (1.00, 0.04 to 1.96). We conducted “leave one out” sensitivity analyses, ^25^ which tested the effect of dropping each jurisdiction from meta-analyses. Combined effects for mental health conditions became significant with the exclusion of both Comcare (-1.74, -3.41 to -0.08) and South Australia (-1.71, -3.42 to 0.00), and for musculoskeletal conditions with the exclusion of the Northern Territory (-0.69, -1.35 to -0.03). Other trauma claims remained non-significant. These results are presented in Supplementary Figure 6.

## Discussion

### Interpretations of step-down effects on scheme exit rates

The local effect of step-downs on scheme exit rates was negative. The first potential explanation is that step-downs reduce the likelihood of return to work. This seems implausible given its lack of theoretical coherence and evidence that greater benefit generosity is positively associated with claim duration.^10,35^

The second interpretation is that step-downs have an anticipatory effect, where claimants leave the compensation system early to avoid reductions in income. As evidence for this interpretation, regression discontinuity plots suggest that where effects were statistically significant, scheme exit rates increased in the week prior to step-down.

An alternative explanation posits that we mis-specified step-downs as occurring earlier than they actually do. This would be the result of our use of cumulative determinations of when step-downs apply contrary to jurisdictions that use calendar determinations, leading to discrepancies. For instance, in Victoria and South Australia, a claimant who works one day in a five-day workweek would be subjected to a step-down after 13 weeks. In our dataset, this would correspond to 13 days or 2.6 weeks of compensated time off work and we would not count them as being affected by step-downs. However, we have identified several reasons to reject these discrepancies as the driver of the negative effect. For one, there were significant anticipatory effects in Comcare, where step-downs are determined by cumulative compensated time off work,^4^ as in our determination. For another, divergent estimates would be attributable to failed return to work attempts and graduated/partial working arrangements. Such claimants have demonstrated positive action to return to work and financial incentives may not provide a sufficient motivation to achieve sustained return to work. Additionally, claimants with graduated/partial working arrangements are less affected by step-downs since only the compensated portion of their wages are reduced. In Comcare, step-downs magnitudes decrease for claimants with partial working arrangements.^4^ We would also expect such exits to be more evenly distributed prior to step-downs. Instead, as noted above RDD plots suggest they are clustered in the week prior to step-down in a manner that deviates from the secular trend. This suggests these claimants are maximising payments under the higher initial rate of compensation.

Our analytical approach – the regression discontinuity design – can only test local effects, i.e., at the cut-off. Evidence that greater benefit generosity increases time off work^10,35^ suggests step-downs may still have longer-term effects, even where there are no local effects. Plotted exit rate patterns generally indicate continuing logarithmic decay, particularly where local effects were non-significant. While this does not rule out longer-term effects, it suggests they are at most relatively small.

### Heterogeneity of effects and potential causes

A moderate proportion of the variance in effects was attributable to heterogeneity. While neither the timing nor magnitude of step-downs were a significant moderator, there were only eight data points for the meta-regression, limiting statistical power. These factors may yet explain some of the differences in effect. For instance, most significant effects were observed among step-downs occurring at 13 weeks, the earliest timing. This aligns with employer and policymaker opinion that delaying step-downs diminishes their effectiveness.^4,5^ However, the 45-week Comcare step-down, the latest tested in this study, also had a significant effect. This suggests unmeasured factors such as the presence of organised unions, who can warn claimants about impending step-downs, may modify step-down effects, regardless of timing.

### Effects by type of injury

There were significant combined effects in fracture claims and more tenuous evidence for effects in mental health and musculoskeletal condition claims. Fractures are generally considered less responsive to benefit generosity since they are more visible and easier to diagnose^35^ with less variability in recovery time.^36^ In other words, there is less discretionary time off work that may be influenced by benefits. Though contrary to expectations, the findings are not unprecedented. We previously found time off work among fracture claims sharply increased after Victoria raised the maximum wage replacement cap from 150% to 200% of average state earnings.^10^ This may be explained by the subset of fracture claims exposed to step-downs. Supplementary Figure 2 illustrates that unlike other injuries, fracture exit rates peak around two months post-claim, possibly reflecting the natural course of recovery.^36^ Claims exceeding this peak will be more complex on average and may be more responsive to benefit generosity.^37^

Mental health conditions are less visible and harder-to-diagnose, characteristics thought to increase sensitivity to benefit generosity. To our knowledge, our previous work is the only empirical investigation of how such claims respond to rate of compensation, though we found no evidence of an effect.^10^ However, the previous study examined the effect of initial rates of compensation, while here we measure a change in that rate. The psychological vulnerability of mental health claimants may mean the act of cutting benefits has a greater effect on scheme exit than variations in what they are paid from the start.

Musculoskeletal conditions are similarly less visible and harder-to-diagnose, with a substantial body of literature demonstrating sensitivity to benefit generosity.^35^ The findings for other trauma were non-significant, though it would be premature to dismiss this as no effect given the combined point estimate was the largest in magnitude. Null results do not necessarily entail null effects.

### Statistical versus practical significance of findings

While the findings were statistically significant, practical significance is less clear. For one, effects were fairly small. At the state level, the largest effect was -2.24 in South Australia. At injury level, the biggest effect was -5.28 among mental health claims in South Australia, though this and the other larger injury effect estimates had wide confidence intervals. Nevertheless, if these are reflective of the maximum potential impact of step-downs, they remain marginal. And if they are indeed anticipatory, the effects may be short-lived, with scheme exit rates returning to normal shortly after step-downs apply.

Step-downs may have negative side effects on claimants. They have been linked to financial strain,^6,38^ which could worsen outcomes or even delay scheme exit, particularly later in the process.^37^ Further, economically-motivated return to work such as that driven by compensation benefits can increase the likelihood of reinjury.^39^

Scheme exit does not necessarily entail return to work and may result in cost-shifting to other income replacement systems.^8,40^ However, it seems unlikely that those who leave workers’ compensation in response to step-downs would go elsewhere if the causal mechanism is financial pressure. Other government-provided incapacity benefits are less generous than workers’ compensation.^3,40^ Some claimants may retire as this option generally entails less financial stress than other.^41^ However, these inferences assume an informed, calculated, and rational economic response to financial incentives. The cut in benefits may induce a negative psychological reaction in some claimants and lead to a scheme exit that is neither return to work nor an alternative that improves financial well-being.

Meta-analyses suggested there was a moderate amount of heterogeneity between jurisdictions, which makes it difficult to make inferences about generalisability. However, the effects varied from small to approximately null, with a positive effect in a single subgroup (musculoskeletal conditions in the Northern Territory). The findings may be applicable to similarly cause-based, devolved workers’ compensation systems in developed economies like Canada and the United States, or other disability-based systems in developed countries, though it is unclear what the effects may be in underdeveloped settings.

### Strengths and limitations

This study has several limitations, some we have already mentioned including discrepancies in determination of step-downs and inconsistent application of step-downs for some claimants. Regression discontinuity designs assume populations around the cut-off are unable to manipulate what condition they are exposed to.^19^ Our study inverted this assumption, since claimants were reacting to the step-down cut-off rather than being allocated by it to separate conditions. The theoretical implications are unclear, though it may provide greater flexibility in interpretation. Rather than simulating a randomised controlled trial as is the case when regression discontinuities meet certain assumptions,^20^ we can interpret the findings more qualitatively.^21^ Similar natural experiment designs like the interrupted time series also consider anticipatory effects.^42^ However, this means we also lose some of the strength in making causal attributions that a simulated randomised controlled trial would provide.

This study also has several strengths. We applied a robust quasi-experimental approach, the regression discontinuity design, to national workers’ compensation data with population-level coverage. There were sufficient data to investigate impact by jurisdiction and most injury subgroups, and meta-analysis increased precision of estimates and provided evidence that effects varied by jurisdiction. Sensitivity analyses provided evidence that effects were not attributable to co-occurring incentives that may have confounded results.

### Conclusions

The findings suggest that step-downs have an anticipatory effect, leading some workers’ compensation recipients to leave the system early in anticipation of a reduction in income. However, the effects are small and probably short-lived. Step-downs may still reduce costs to workers’ compensation systems, which is a legitimate policy goal. However, our findings suggest step-downs have marginal practical significance and are generally ineffective as a return to work policy initiative.

### Ethics

This study received ethics approval from the Monash University Human Research Ethics Committee (CF14/2995 – 2014001663).

## Data Availability

Aggregated data are publicly available on a FigShare repository, along with analytical code. Case-level data contain identifiable information and have not made available. We have provided cleaning file data at the FigShare repository to demonstrate our approach. The data source is the National Data Set for Compensation-based Statistics, provided by Safe Work Australia

https://doi.org/10.26180/5dba1e5b4277a

## Acknowledgements

We would like to thank the COMPARE Project’s Advisory Group who provided valuable insight when interpreting the results. We would also like to thank the two anonymous peer reviewers who provided invaluable commentary and greatly improved the quality of the manuscript.

---

**Supplementary Figure 1.**
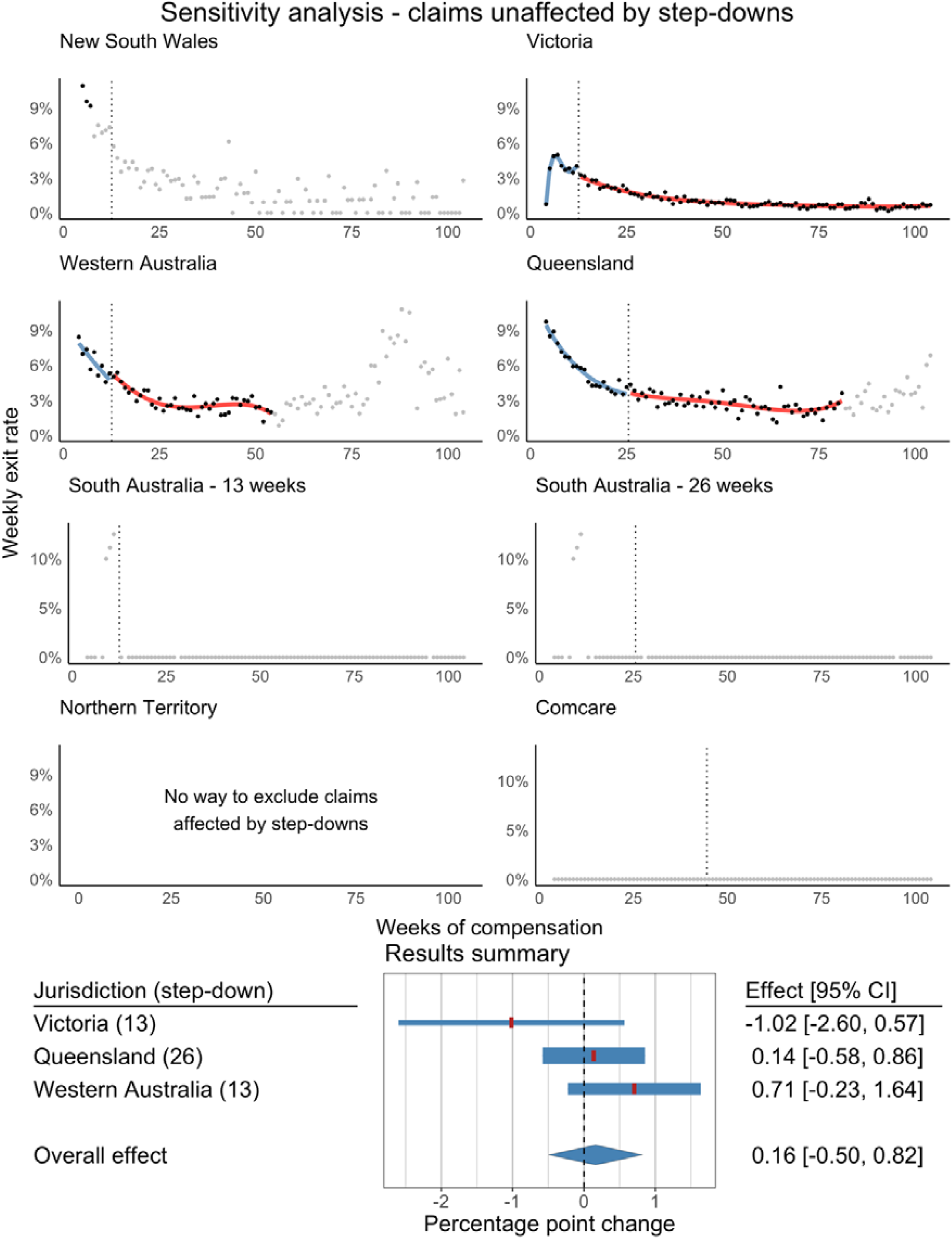
Sensitivity analysis - claims unaffected by step-downs. Regression discontinuity plots and meta-analysis.

**Supplementary Figure 2.**
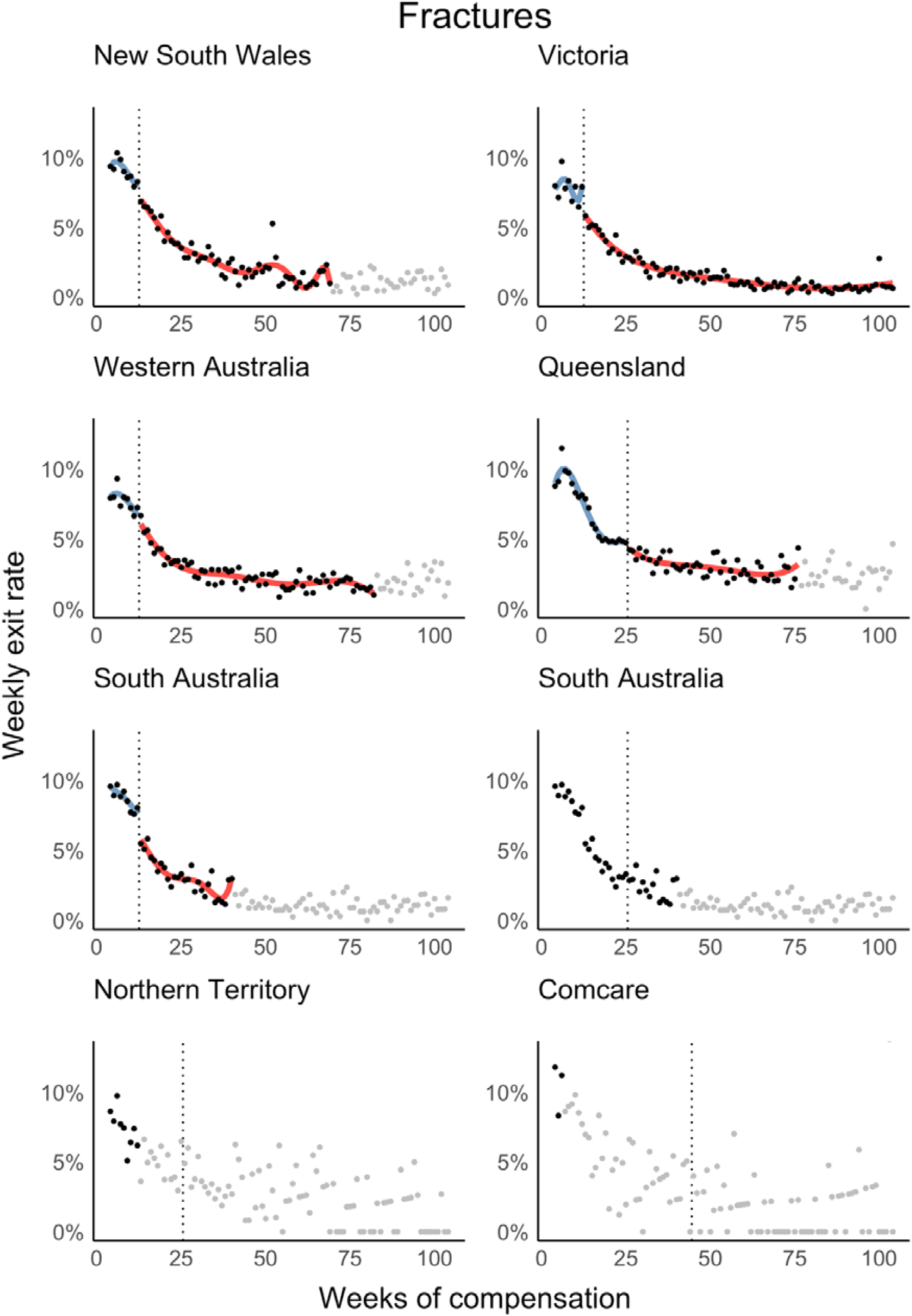
Fracture subgroup regression discontinuity plots.

**Supplementary Figure 3.**
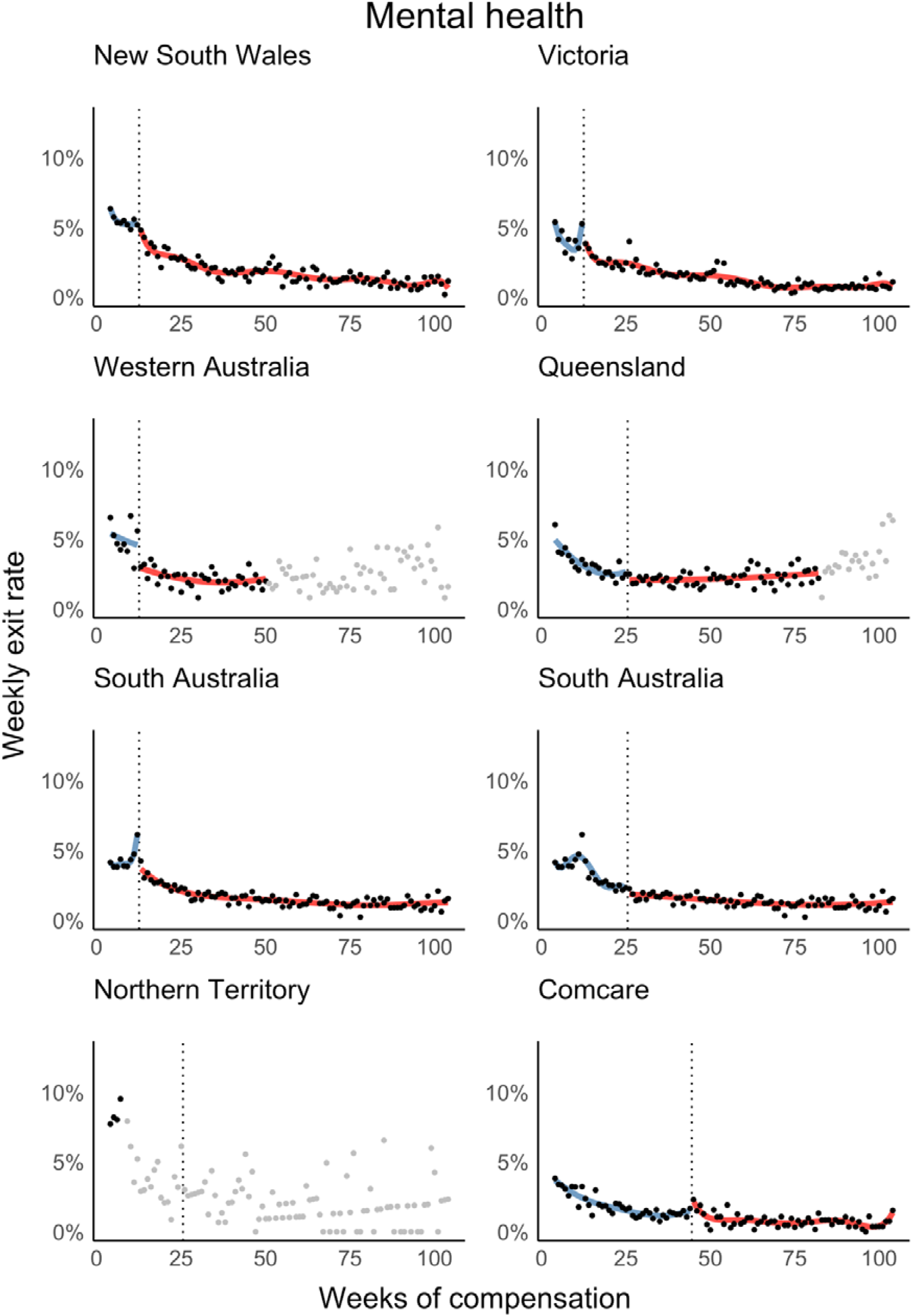
Mental health regression discontinuity plots.

**Supplementary Figure 4.**
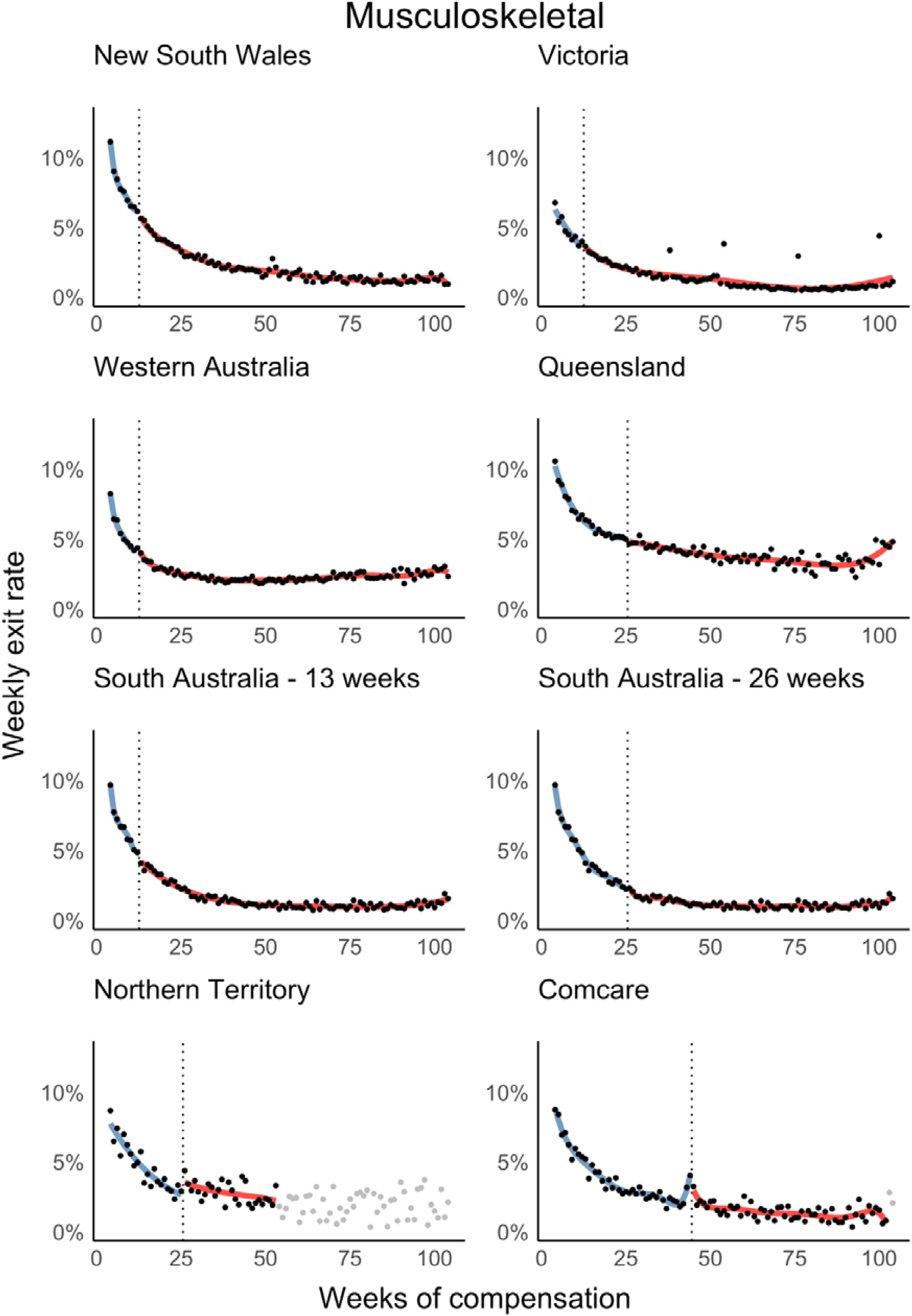
Musculoskeletal regression discontinuity plots.

**Supplementary Figure 5.**
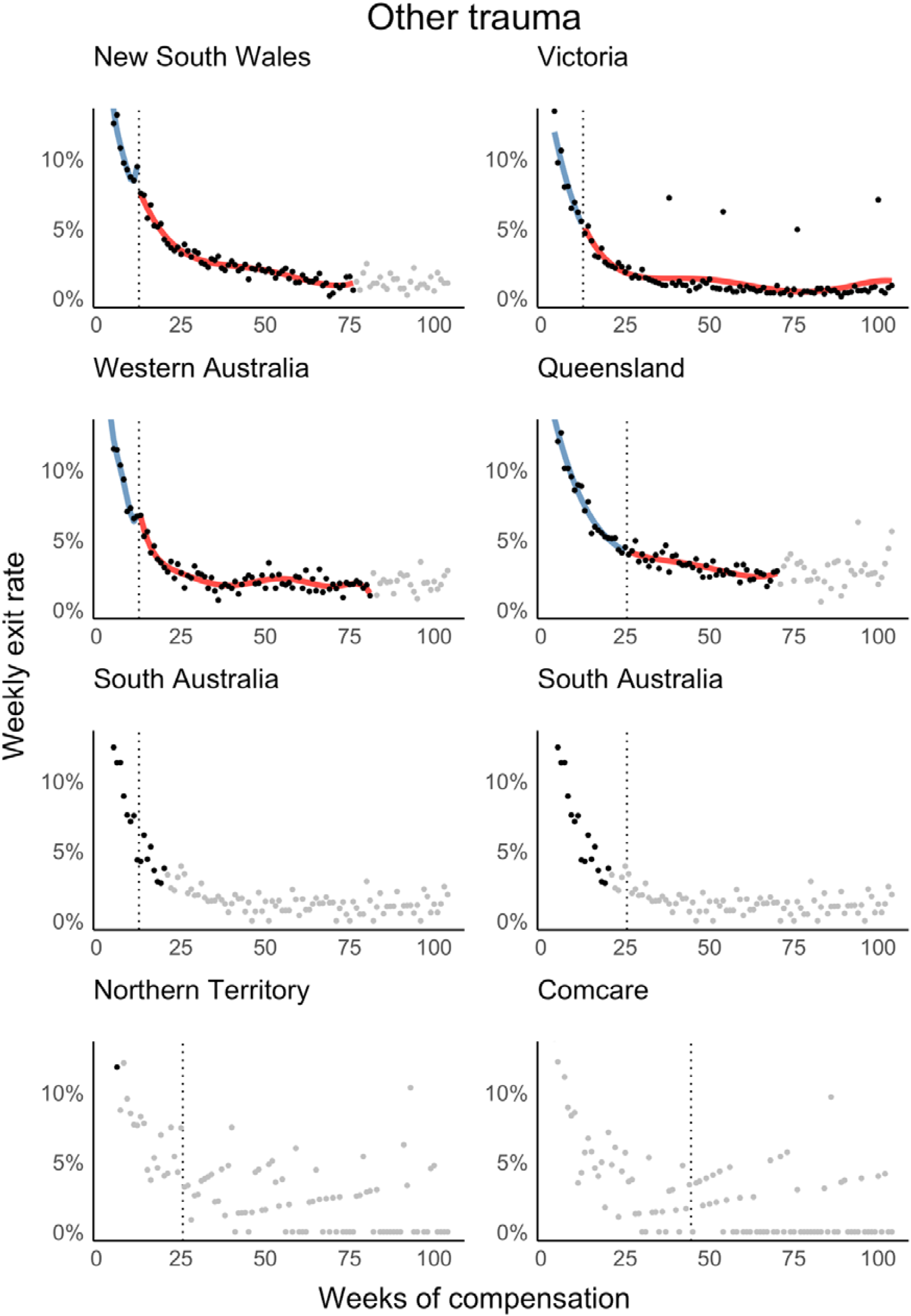
Other trauma regression discontinuity plots.

**Supplementary Figure 6.**
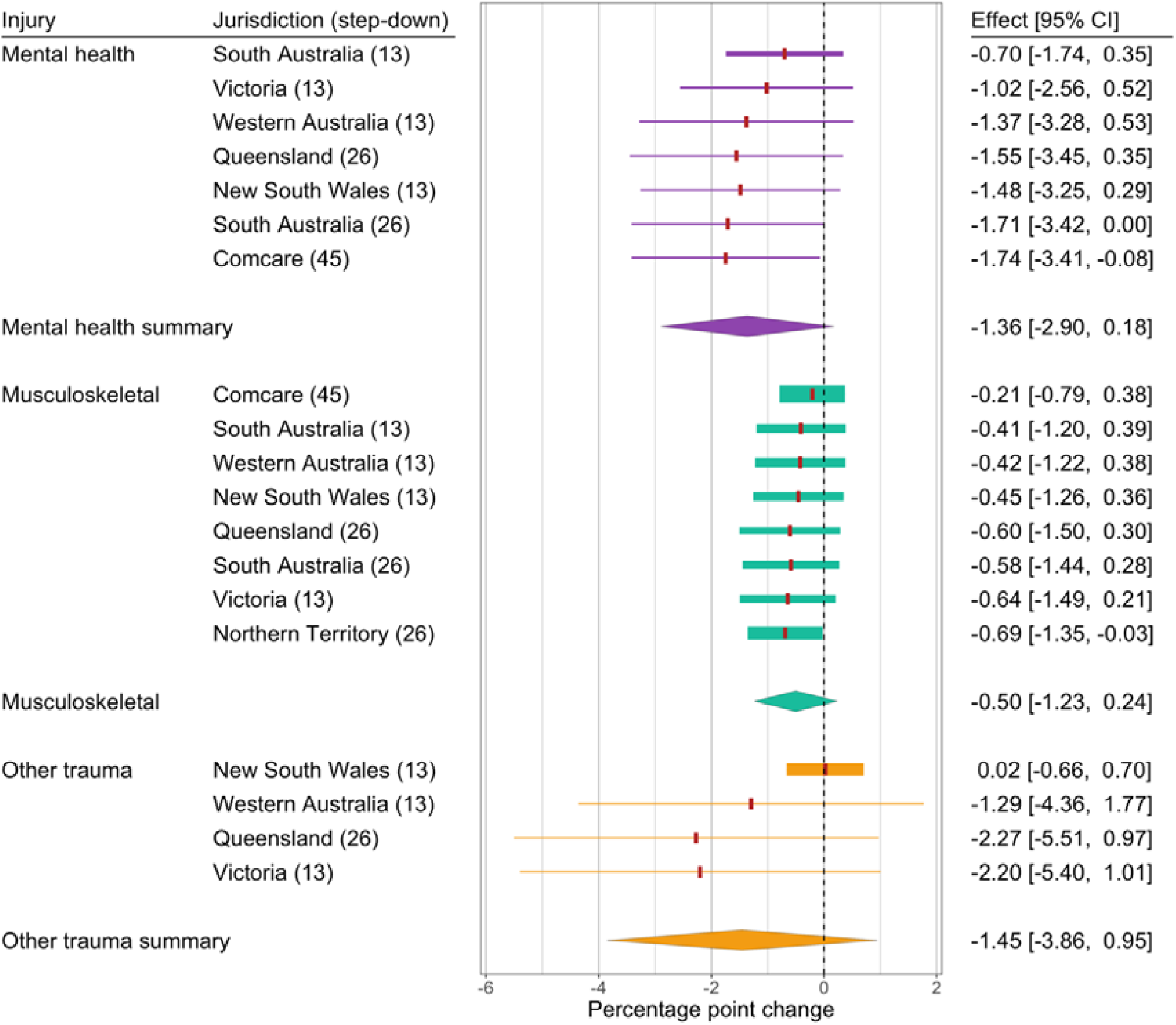
Sensitivity analysis – “leave one out” meta-analysis by injury subgroup.

